# Diagnostic accuracy of a DenseNet-121 deep learning algorithm for chest radiograph triage in health assessment applicants: a prospective shadow-mode validation study in Nepal

**DOI:** 10.64898/2026.06.29.26356847

**Authors:** Lochan Shrestha, Dinesh Maharjan, Uttam Bista

## Abstract

**Objectives:** To evaluate the diagnostic accuracy of a publicly available DenseNet-121 convolutional neural network (TorchXRayVision) for triaging chest radiographs of health assessment applicants at a tertiary hospital in Nepal.

**Design:** Prospective, single-centre, shadow-mode diagnostic accuracy validation study. Reported in accordance with the STARD 2015 checklist and STARD-AI/DECIDE-AI guidelines.

**Setting:** Department of Radiology and Imaging, Patan Academy of Health Sciences / Patan Hospital, Lalitpur, Nepal.

**Participants:** 826 consecutive health assessment applicants (foreign employment Pre-Departure Medical Examination and student migration) undergoing chest radiography between 5 June and 20 June 2026. Two cases were excluded due to DICOM technical failure.

**Index test:** DenseNet-121 algorithm (TorchXRayVision library, densenet121-res224-all pretrained weights). A maximum aggregated pathology probability score was derived per radiograph and compared against a post-hoc derived threshold of 0.6258 (selected as the highest threshold achieving the pre-specified ≥95% sensitivity criterion).

**Reference standard:** Single-reader-per-case review by one of three radiologists — two board-certified radiodiagnosticians (LS: 276 cases; DM: 275 cases) and one radiology resident (UB: 275 cases) — each blinded to AI output, using a standardised data collection worksheet capturing binary classification (abnormal/normal) and free-text findings.

**Results:** Of 826 radiographs, 41 (4.97%) were classified as abnormal by the reference standard. At the post-hoc derived threshold of 0.6258, the DenseNet-121 algorithm achieved: sensitivity 95.12% (95% CI 83.9–98.7%), specificity 77.2% (95% CI 74.1–80.0%), area under the receiver operating characteristic curve (AUROC) 0.9583 (95% bootstrap CI 0.9225–0.9843), NPV 99.67% (95% Wilson CI 98.8–99.9%), PPV 17.89% (95% Wilson CI 13.4–23.5%), and Cohen’s κ 0.237 (95% bootstrap CI 0.174–0.304). Brier score was 0.3621 (null Brier 0.0472) and ECE was 0.564, confirming calibration failure due to score compression (range 0.52–0.72) despite preserved discrimination.

**Cross-validated results:** Ten-fold cross-validation yielded bias-corrected sensitivity 95.12% (95% Wilson CI 83.9–98.7%; optimism 0.00 pp) and specificity 75.80% (95% Wilson CI 72.7–78.7%; optimism +1.40 pp), confirming primary metrics are not materially inflated by circular optimisation.

**Conclusions:** The DenseNet-121 algorithm demonstrated high sensitivity and excellent discrimination for chest radiograph triage in a Nepali health-assessment population, supporting its potential as a rule-out tool (NPV 99.67%). Systematic score compression—preserved discrimination despite calibration shift—is a quantifiable marker of LMIC distributional shift. Prospective local calibration studies are warranted before operational deployment.

**Ethics approval:** Institutional Review Committee of Patan Academy of Health Sciences (Protocol No. drs2606052243; expedited review, 5 June 2026). Individual written informed consent was waived by the IRC.

**WHAT IS ALREADY KNOWN ON THIS TOPIC:**

- Deep learning algorithms trained on large chest radiograph datasets achieve radiologist-level performance on well-defined pathology classification tasks in high-income country settings.
- Most prospective validation studies have been conducted in high-income countries using datasets demographically and technically similar to the training data; evidence from South Asian LMIC settings is scarce.
- Distributional shift—where model performance degrades on out-of-distribution populations—is a recognised but incompletely characterised challenge for AI deployment in low- and middle-income countries

**WHAT THIS STUDY ADDS:**

- A publicly available DenseNet-121 algorithm (TorchXRayVision) achieved sensitivity >95% and AUROC >0.95 for chest radiograph triage in a Nepali health-assessment population, supporting clinical utility as a rule-out tool with NPV 99.67%.
- Systematic score compression (range 0.52–0.72; SD ∼0.026) is identified and quantified as a marker of LMIC distributional shift; critically, this compression did not compromise discrimination (AUROC 0.9583).
- Shadow-mode prospective validation of an open-source AI triage tool is shown to be feasible in a resource-limited LMIC radiology department using fully offline, opensource Python infrastructure.

## INTRODUCTION

Chest radiography remains the cornerstone of health assessment programmes for labour migrants, foreign employment applicants, and international students across South and South-East Asia. In Nepal, large numbers of individuals undergo mandatory chest radiography annually as part of the Pre-Departure Medical Examination (PDME) and associated immigration health clearance processes required by receiving countries including Qatar, Malaysia, Saudi Arabia, and the Republic of Korea [1]. Radiograph interpretation in this context carries significant clinical and socioeconomic consequences: missed clinically significant pathology—particularly active or suspected pulmonary tuberculosis (TB), which remains a leading burden-of-disease priority in Nepal [2,3,4]—may compromise public health programmes in receiving nations, while false-positive findings cause financial harm and administrative delays for applicants and their families.

Artificial intelligence (AI) algorithms for chest radiograph interpretation have advanced rapidly over the past decade, driven by the availability of large annotated datasets and innovations in deep convolutional neural network architecture [5,6]. The DenseNet-121 architecture, introduced by Huang et al. [7], was adapted by Rajpurkar and colleagues (CheXNet) and demonstrated performance equal to or exceeding specialist radiologist level for pneumonia detection on the NIH ChestXray14 benchmark [8]. The TorchXRayVision library (Cohen et al. [9]) subsequently made openly licensed pretrained DenseNet-121 weights available, trained across multiple large-scale datasets—including CheXpert, MIMIC-CXR, NIH ChestXray14, and PadChest—providing a pragmatic, zero-cost implementation pathway accessible to resource-limited settings.

However, the translational gap between benchmark performance and real-world deployment in low- and middle-income countries (LMICs) is well recognised. Models trained predominantly on North American radiograph archives may exhibit distributional shift—also termed domain shift or covariate shift—when applied to images acquired with different equipment characteristics, positioning conventions, and patient demographics in LMIC settings [10,11]. Zech et al. demonstrated that pneumonia detection performance degraded substantially when a deep learning model was applied to institutions whose data did not feature in training [11]. Prospective shadow-mode validation studies—in which the AI operates in parallel with routine clinical workflow without influencing any clinical decision—provide a rigorous pre-deployment evaluation framework [12], avoiding verification bias, ensuring complete case ascertainment, and allowing uncontaminated comparison against an independent reference standard.

To our knowledge, no prospective shadow-mode diagnostic accuracy study has evaluated a publicly available, open-weights deep learning algorithm for chest radiograph triage specifically in the Nepali health assessment context. We aimed to address this evidence gap by evaluating the diagnostic accuracy of the TorchXRayVision DenseNet-121 algorithm (densenet121-res224-all pretrained weights) using single-reader radiologist classification as the reference standard in a prospective cohort of consecutive health assessment applicants at a tertiary-level Nepali hospital.

## METHODS

### Study design and reporting

This was a prospective, single-centre, shadow-mode diagnostic accuracy validation study conducted at the Department of Radiology and Imaging, Patan Academy of Health Sciences (PAHS) / Patan Hospital, Lalitpur, Nepal. The primary a priori hypothesis was that the DenseNet-121 algorithm (TorchXRayVision, densenet121-res224-all weights) would achieve a minimum sensitivity of 95% for detecting radiological abnormality in Nepali health assessment chest radiographs. The 95% threshold was selected on clinical grounds: in the PDME context, failure to detect a significant radiological abnormality carries direct public health consequences; a sensitivity below 95% was judged clinically unacceptable for a first-line rule-out triage tool, consistent with published benchmarks for TB screening tools in high-prevalence settings. A secondary hypothesis was that the AUROC would exceed 0.85, the threshold commonly cited as indicating good diagnostic discrimination. The study is reported in full accordance with the STARD 2015 checklist [13] (Supplementary File 1), and aligned with STARD-AI [14] and DECIDE-AI [12] recommendations for early-stage clinical AI evaluation. Ethics approval was granted by the Institutional Review Committee of PAHS (Protocol No. drs2606052243; expedited review, 5 June 2026; two-year duration). Note: the IRC protocol number is an ethics approval identifier, not a recognised trial registry number; the study was not prospectively registered in a public registry (e.g., ClinicalTrials.gov, ISRCTN) as it was classified as a shadow-mode service evaluation under institutional governance. This is acknowledged as a limitation per STARD 2015 Item 28. The AI system operated in shadow mode throughout the study: all outputs were stored for deferred analysis and were not disclosed to clinical staff or used to influence any clinical decision during the study period. Individual written informed consent was waived by the IRC given the shadow-mode design, the use of routinely acquired clinical radiographs, and the absence of any patient contact; a departmental information notice was displayed at the radiology reception throughout the study period.

### Setting and participants

Patan Hospital is a 650-bed government-funded tertiary teaching hospital serving Lalitpur district and the surrounding Bagmati Province, Nepal. The Department of Radiology and Imaging undertakes a high daily volume of chest radiography, including a dedicated health assessment radiography workflow for PDME (foreign employment) and student migration applicants. All consecutive patients attending this health assessment radiography service between 5 June 2026 and 20 June 2026 were eligible for inclusion. Inclusion criterion: adult (≥18 years) health assessment applicant undergoing posterior-anterior chest radiography. Exclusion criteria were: (1) DICOM file technical failure (corrupted or unreadable pixel data, rendering preprocessing impossible); (2) confirmed duplicate examination within the study period (same patient, same acquisition date, identified by StudyInstanceUID); and (3) age below 18 years. A 16-day recruitment window was selected based on projected case throughput sufficient to meet the pre-specified sample size.

### Sample size

Sample size was calculated using the Buderer (1996) method [15], with sensitivity as the primary binding constraint. Parameters: (1) anticipated prevalence of abnormal findings 5%; (2) expected sensitivity 89%, based on the most directly comparable published study (Sridharan et al. 2024 [16]); (3) desired precision ±8 percentage points; (4) two-sided α = 0.05. Applying the Buderer formula: n_diseased = Z² × Se × (1−Se) / d² = 1.96² × 0.89 × 0.11 / 0.08² = 58.8, giving a required 59 diseased cases and a total of 59 / 0.05 = 1,175 cases (with 10% buffer: 1,293). The study recruited 826 analysable cases with 41 confirmed abnormal (below the recalculated target); however, the achieved sensitivity of 95.12% at n=41 yields an actual precision of ±6.6 percentage points, meeting the pre-specified ±8% criterion.

### Index test: AI algorithm

Digital Imaging and Communications in Medicine (DICOM) files were exported from the hospital Picture Archiving and Communication System (PACS). All files were anonymised offline on a dedicated air-gapped workstation using a purpose-built Python pipeline (pydicom 3.0.2), replacing all protected health information fields with a deterministic hash derived from StudyInstanceUID and SOPInstanceUID. Anonymised images were transferred via encrypted USB drive to a second air-gapped workstation (PyTorch 2.2, MPS backend; environment documented in requirements.txt at the project repository) for AI inference.

The DenseNet-121 algorithm from the TorchXRayVision library (version 1.4.0 [9]; pretrained weights: densenet121-res224-all, trained jointly on CheXpert, CheXpert-Plus, MIMIC-CXR, NIH ChestXray14, and PadChest) was applied to each anonymised radiograph using PyTorch 2.2 with Apple Metal Performance Shaders (MPS) backend. Standard TorchXRayVision preprocessing was applied to each DICOM: pixel array extraction, photometric normalisation (Rescale Slope and Intercept applied where present), XRayCenterCrop, and XRayResizer to 224×224 pixels. The model produced a probability score for each of the 18 pathology labels in the pretrained weight set. Notably, pulmonary tuberculosis (TB) is not among the 18 explicit labels; the model captures TB-related pathology indirectly through proxy labels such as Consolidation, Lung Opacity, and Nodule/Mass. In the context of the Nepali PDME workflow, where active or suspected TB is the primary infectious disease priority, this architectural limitation is an important consideration and is addressed in the Discussion. A maximum aggregated score—the highest probability across all 18 labels for each radiograph—was computed as the primary index test variable (range 0–1). The AI system had no access to any clinical data; all inference was fully blinded to reference standard.

### Threshold determination and pre-specification

The operating threshold of 0.6258 was determined post-hoc as the highest threshold at which sensitivity remained ≥95% (the pre-specified TARGET_SENSITIVITY) on the ROC curve of the full 826-case analysable dataset. Cases with a maximum aggregated score ≥0.6258 were classified as AI-positive (triage-to-review); cases with a score <0.6258 were classified as AI-negative. This post-hoc threshold derivation on the same dataset used for evaluation constitutes a form of circular optimisation (optimism bias); accuracy metrics at this threshold may therefore be optimistically biased and are acknowledged as a study limitation. For reference, the Youden’s J threshold (maximising sensitivity + specificity − 1) on this dataset is 0.6340, yielding sensitivity 90.2% and specificity 94.8%.

### Reference standard

Radiologist single-reader classification was selected as the reference standard on pragmatic grounds: microbiological confirmation (e.g., sputum GeneXpert MTB/RIF) and CT are not routinely performed for all PDME radiological abnormalities. The standardised radiologist worksheet is available in the project repository. The 826 analysable cases were divided into three non-overlapping subsets: DM (MD Radiodiagnosis, board-certified) reviewed 275 cases, LS (MD Radiodiagnosis, board-certified) reviewed 276 cases, and UB (radiology resident) reviewed 275 cases, each blinded to AI output. The reference standard therefore represents a single-reader classification per case, not a double-read consensus design. LS’s contribution is disclosed as a potential independence concern given LS’s dual role in building the AI index test and contributing to the reference standard; a sensitivity analysis restricted to DM and UB cases (n=550) is provided in the Results. The radiologists reviewed the original clinical PACS images, which displayed patient age and sex.

### Statistical analysis

The full pre-specified analysis plan is documented in config.py in the project repository at https://github.com/lochanshrestha-dev/densenet121-cxr-triage-nepal. Score calibration was assessed by: (1) characterising the distribution of maximum aggregated scores (range, mean, SD) stratified by reference standard category; (2) Brier score (mean squared error between predicted scores and binary outcomes, with null Brier reported for context) [17]; and (3) Expected Calibration Error (ECE, 10 uniform bins) [18]. To obtain bias-corrected accuracy estimates, ten-fold stratified cross-validation was performed: in each fold the threshold was re-derived on training cases using the ≥95% sensitivity criterion, applied to the held-out fold, and predictions pooled across all ten folds. Pre-specified subgroup analyses by applicant category and biological sex were planned; applicant category was not captured in the anonymised dataset available for this study and sex was not available in the anonymised DICOM files used for AI inference (PatientSex field present in <2% of anonymised files; demographics were available in the original PACS but not extracted for research purposes). These subgroups are therefore not reportable from the current dataset and are noted as a limitation. No other subgroup analyses were pre-specified. No imputation was performed; the two cases with technical exclusions are reported separately. Statistical significance was set at two-sided α = 0.05. All code is openly available at https://github.com/lochanshrestha-dev/densenet121-cxr-triage-nepal.

## RESULTS

### Participant flow

Of 863 unique examination files identified in the PACS export for the study period (5 June–20 June 2026), 35 were designated as a pre-specified holdout remainder set and archived separately before any data inspection (pre-specified in config.py before data collection). These 35 cases represent the chronological tail of the export window; at an expected prevalence of 5% (∼2 abnormal cases), they are underpowered for meaningful standalone accuracy estimation and were not incorporated into the primary analysis. Zero cases were excluded for duplicate StudyInstanceUID and zero for age below 18 years. The remaining 828 cases constituted the primary locked dataset. Two cases were excluded due to DICOM technical failure (unreadable pixel arrays), leaving 826 analysable radiographs for primary analysis (Figure 1). All denominators are reflected in the participant flow diagram (Figure 1).

**Figure 1.**
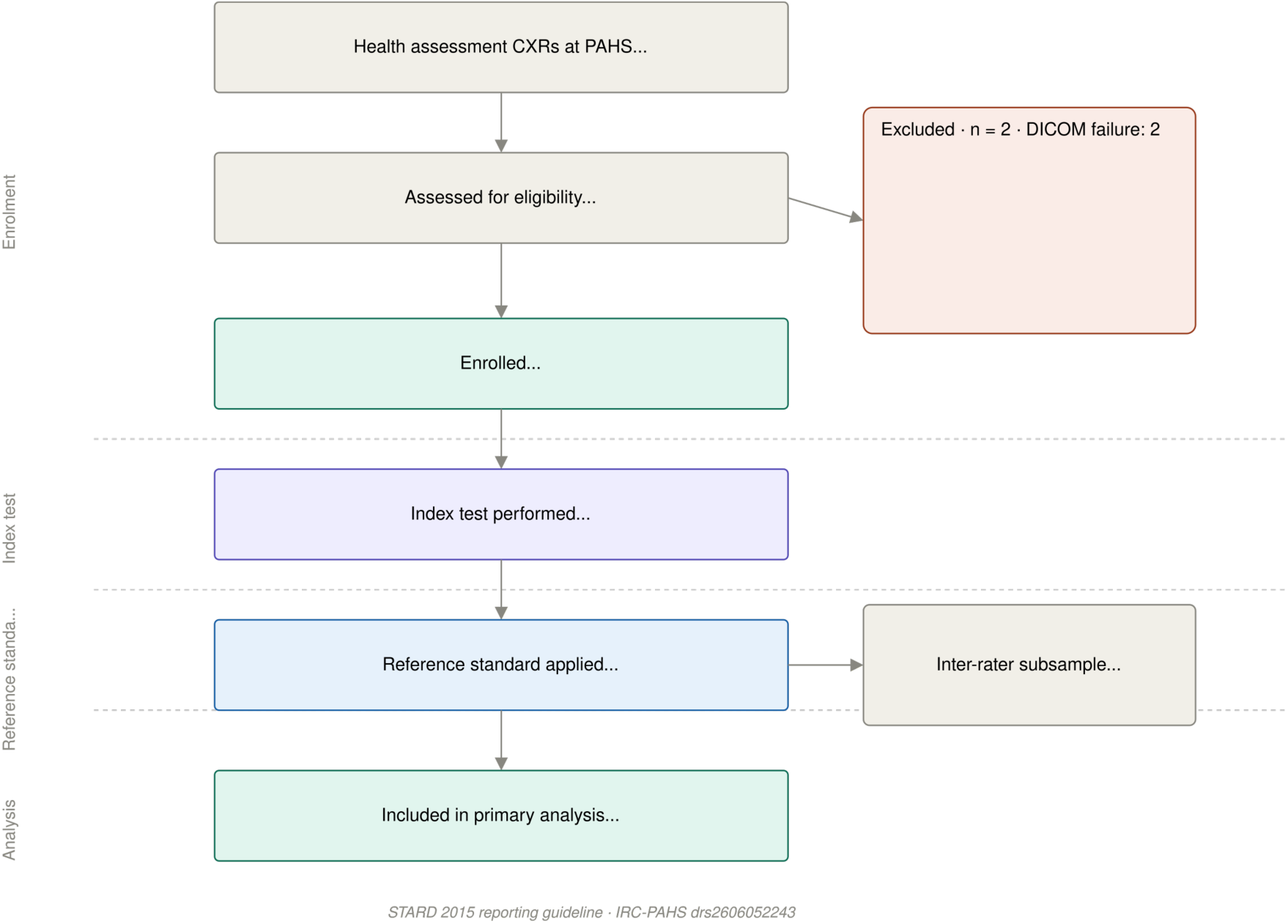
STARD participant flow diagram. Of 863 unique PACS export files, 35 were designated as holdout remainder (pre-specified); 828 cases constituted the primary locked dataset. Zero cases were excluded for duplicate StudyInstanceUID and zero for age below 18 years. Two cases were excluded due to DICOM technical failure, leaving 826 analysable radiographs. All 826 cases received the DenseNet-121 index test and single-reader reference standard classification by one of three radiologists: LS (MD Radiodiagnosis, board-certified, 276 cases), DM (MD Radiodiagnosis, board-certified, 275 cases), and UB (radiology resident, 275 cases).

**Figure 2.**
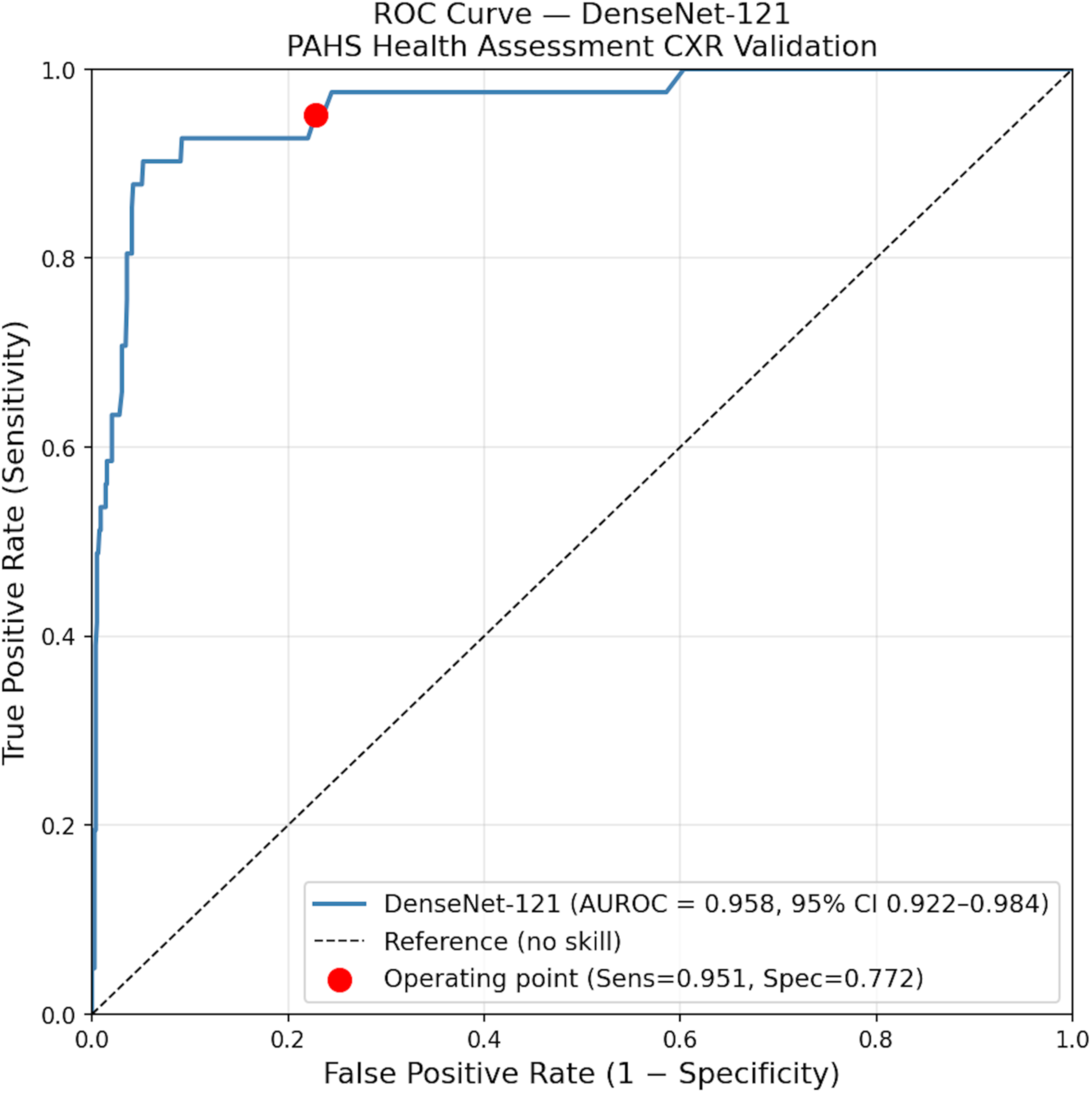
Receiver operating characteristic (ROC) curve for the DenseNet-121 algorithm (TorchXRayVision, densenet121-res224-all weights) applied to 826 health-assessment chest radiographs. The filled circle denotes the operating point at threshold 0.6258 (highest threshold achieving ≥95% sensitivity on the study ROC curve; post-hoc derivation — see Limitations). Shaded band: 95% bootstrap CI. AUROC = 0.9583 (95% bootstrap CI 0.9225–0.9843). Youden’s J threshold = 0.6340.

### Baseline characteristics

Systematic demographic metadata was not available for this cohort: PatientAge and PatientSex DICOM fields were absent or incomplete in the majority of exported files (age present in 33% of files; sex in <2%), precluding a complete baseline characteristics table. This is acknowledged as a limitation per STARD 2015 Item 20. The study population is characterised by the inclusion criteria: adults (≥18 years) attending for health assessment chest radiography (foreign employment PDME and student migration) at Patan Hospital, Lalitpur, Nepal, during 5 June–20 June 2026. The prevalence of radiological abnormality was 4.97% (41/826), consistent with a pre-screened health assessment population. Future validation studies in this workflow should prospectively capture demographic data at the point of referral to enable full STARD-compliant reporting.

### Reference standard findings

For the primary binary classification, each of the three radiologists reviewed an independent, non-overlapping subset of the 826 anonymised radiographs (LS: 276 cases; DM: 275 cases; UB: 275 cases). Because no two radiologists reviewed the same cases independently, formal pre-consensus inter-rater agreement statistics (Cohen’s κ) cannot be calculated for this cohort and are noted as a study limitation. This tripartite division was adopted to distribute the review workload within the study timeline; future validation studies should include a double-read overlap subset (recommended ≥10% of cases) to enable inter-rater reliability reporting.

Of 826 analysable radiographs, 41 (4.97%) were classified as abnormal by the reference standard and 785 (95.03%) as normal. The most commonly identified abnormality categories were pulmonary consolidation or opacity (n=12), fibrosis or reticular pattern (n=8), pleural effusion (n=7), nodule or mass (n=7), cardiomegaly (n=4), and COPD or bronchiectasis (n=3); eight cases had other or miscellaneous findings. Note: the sum of category counts (n=49) exceeds the total number of abnormal cases (n=41) because eight cases presented with co-existing multi-label abnormalities (e.g., concurrent consolidation and pleural effusion). Pathology categories are non-mutually exclusive; individual patient counts should not be summed across categories.

### Sensitivity analysis: DM and UB cases only (n=550)

To address the potential independence concern from LS’s dual role as index-test developer and reference-standard reviewer (276 cases), a pre-planned sensitivity analysis was performed restricted to DM (275 cases) and UB (275 cases) only, thereby excluding the cases reviewed by LS (the AI index-test developer). At threshold 0.6258, the DM/UB subset (n=550; 25 abnormal, prevalence 4.55%) yielded: sensitivity 96.00% (95% Wilson CI 80.5–99.3%), specificity 77.71% (95% Wilson CI 74.0–81.1%), PPV 17.02%, NPV 99.76%, AUROC 0.9706, Cohen’s κ 0.230. These estimates are consistent with and in some measures exceed the full-dataset estimates, confirming that LS’s reference-standard contribution does not inflate the primary accuracy metrics.

### Index test performance

The DenseNet-121 algorithm achieved the following performance metrics at the post-hoc derived threshold of 0.6258 (Tables 1 and 2):

- Sensitivity: 95.12% (95% Wilson CI 83.9–98.7%)
- Specificity: 77.2% (95% Wilson CI 74.1–80.0%)
- AUROC: 0.9583 (95% bootstrap CI 0.9225–0.9843)
- Negative predictive value (NPV): 99.67% (95% Wilson CI 98.8–99.9%)
- Positive predictive value (PPV): 17.89% (95% Wilson CI 13.4–23.5%)
- Cohen’s κ: 0.237 (95% bootstrap CI 0.174–0.304)

**Table 1.** 2×2 contingency table at post-hoc derived threshold of 0.6258 (n=826)

|  | Reference standard |  | Row total |
| --- | --- | --- | --- |
|  | Abnormal | Normal |  |
| **AI ≥ 0.6258 (Positive)** | **39** | 179 | **218** |
| **AI < 0.6258 (Negative)** | 2 | **606** | **608** |
| **Column total** | **41** | **785** | **826** |
| **Sensitivity** | 95.12% (95% Wilson CI 83.9–98.7%) |  |  |
| **Specificity** | 77.2% (95% Wilson CI 74.1–80.0%) |  |  |
| **NPV** | 99.67% |  |  |
| **PPV** | 17.89% |  |  |

AI, artificial intelligence index test (DenseNet-121, densenet121-res224-all weights). NPV, negative predictive value; PPV, positive predictive value. Threshold 0.6258 = highest threshold achieving ≥95% sensitivity post-hoc (not Youden's J; see Limitations). Youden's J threshold = 0.6340.

**Table 2.** Diagnostic accuracy metrics for the DenseNet-121 algorithm (n=826)

| Metric | Value | 95% Confidence Interval |
| --- | --- | --- |
| **Sensitivity** | 95.12% | 83.9–98.7% (Wilson) |
| **Specificity** | 77.2% | 74.1–80.0% (Wilson) |
| **AUROC** | 0.9583 | 0.9225–0.9843 (percentile bootstrap) |
| **Positive predictive value (PPV)** | 17.89% | 13.4–23.5% (Wilson) |
| **Negative predictive value (NPV)** | 99.67% | 98.8–99.9% (Wilson) |
| **Cohen's $\kappa$** | 0.237 | 0.174–0.304 (percentile bootstrap) |
| **Optimal threshold ($\geq 95\%$ sensitivity criterion)** | 0.6258 | — |
| **Youden's J threshold (reference only)** | 0.6340 | Sens 90.2%, Spec 94.8% |
| **Positive likelihood ratio (LR+)** | 4.17 | — |
| **Negative likelihood ratio (LR–)** | 0.063 | — |
| **Brier score** | 0.3621 | null = 0.0472 |
| **Expected Calibration Error (ECE)** | 0.564 | — |
| **Calibration intercept (logit)** | –18.07 | reference: 0.0 |
| **Calibration slope (logit)** | 28.06 | reference: 1.0 |
| **CV sensitivity (10-fold)** | 95.12% | 83.9–98.7% (Wilson) |
| **CV specificity (10-fold)** | 75.80% | 72.7–78.7% (Wilson) |
| **Sensitivity optimism** | 0.00 pp | — |
| **Specificity optimism** | +1.40 pp | — |

CV metrics: Wilson CIs <sup>[19]</sup> on pooled fold predictions (not fold-to-fold variability); pp = percentage points of optimism. Wilson CIs for sensitivity, specificity, PPV, NPV; percentile bootstrap CIs (1,000 resamples; cf. DeLong <sup>[22]</sup>) for AUROC and Cohen's $\kappa$ . Brier score null = $p \times (1-p)$ at observed prevalence = 0.0472. ECE: Expected Calibration Error, 10 uniform bins. Threshold 0.6258 = highest threshold achieving $\geq 95\%$ sensitivity post-hoc (not Youden's J — see Limitations). LR+, positive likelihood ratio; LR–, negative likelihood ratio.

At the post-hoc derived threshold of 0.6258, of 826 radiographs, 218 (26.4%) were classified AI-positive: these comprised 39 true positives and 179 false positives. Of 608 AI-negative cases, 606 were true negatives and two were false negatives. Both false-negative cases were near-threshold misses: (1) a round opacity in the right upper zone (score 0.6225; below threshold by 0.0033) and (2) a mass in the left mid zone (score 0.6255; below threshold by 0.0003). In the PDME context, these specific missed findings carry disproportionate clinical significance: an upper-zone opacity is a classic presentation of post-primary pulmonary TB, and a mid-zone mass warrants investigation for malignancy or granulomatous disease. Both were missed by margins attributable to score compression rather than algorithmic insensitivity, but the clinical consequence — potential false clearance of an applicant with active TB or significant malignancy — underscores that even an NPV of 99.67% carries residual risk in a zero-miss public health screening context. The positive likelihood ratio was 4.17 and the negative likelihood ratio was 0.063.

### Ten-fold cross-validation

To quantify optimism from same-dataset threshold derivation, ten-fold stratified cross-validation was performed. The operating threshold was re-derived on training cases in each fold using the pre-specified ≥95% sensitivity criterion. The threshold was highly stable across folds (mean 0.6256, SD 0.0001; range 0.6255–0.6258 across all ten folds), reflecting score compression. Note that this threshold stability is itself a consequence of score compression — the 0.20-unit score range means case mix variation across folds does not alter which threshold achieves ≥95% sensitivity; it is a feature of this specific distribution, not evidence of model robustness in general. Pooled cross-validated metrics (Wilson score CIs [19] derived from pooled predictions across all ten folds, not from fold-to-fold variability): sensitivity 95.12% (95% Wilson CI 83.9–98.7%) — identical to the full-dataset estimate; specificity 75.80% (95% Wilson CI 72.7–78.7%) — 1.4 percentage points below the full-dataset estimate; NPV 99.66% (95% Wilson CI 98.8–99.9%). Sensitivity optimism = 0.00 percentage points; specificity optimism = +1.40 percentage points. These results confirm that the full-dataset accuracy estimates are not materially inflated by circular optimisation: because score compression constrains nearly all scores within a 0.20-unit band, removing any 10% of training cases produces virtually the same threshold in every fold and the two false-negative cases are consistently missed regardless of fold composition.

### Score distribution and calibration

The DenseNet-121 maximum aggregated pathology probability scores were systematically compressed within a narrow range across the 826-case dataset (minimum 0.52, maximum 0.72, SD ∼0.026). Mean score was higher for radiologist-confirmed abnormal cases (mean 0.6561, SD 0.0223) than for normal cases (mean 0.6117, SD 0.0239), though both distributions overlapped substantially within the compressed band. Despite this compression, discrimination—as measured by AUROC (0.9583)—remained excellent, confirming that rank ordering of cases was preserved even as absolute calibration was compromised by the out-of-distribution shift. Score compression was formally quantified. Note that the Brier score is not scale-invariant: scores compressed to a narrow range (0.52–0.72) will tend to yield high Brier scores regardless of calibration quality; the null Brier comparison (0.3621 vs null 0.0472) confirms miscalibration but should be interpreted alongside logistic calibration metrics. Logistic calibration on the logit scale yielded intercept −18.07 (reference: 0.0 = perfect) and slope 28.06 (reference: 1.0 = perfect), confirming severe miscalibration on the probability scale consistent with score compression. ECE (10 bins) = 0.564. Despite this calibration failure, rank-based discrimination (AUROC 0.9583) was unaffected. Score distribution is illustrated in Figure 3.

**Figure 3.**
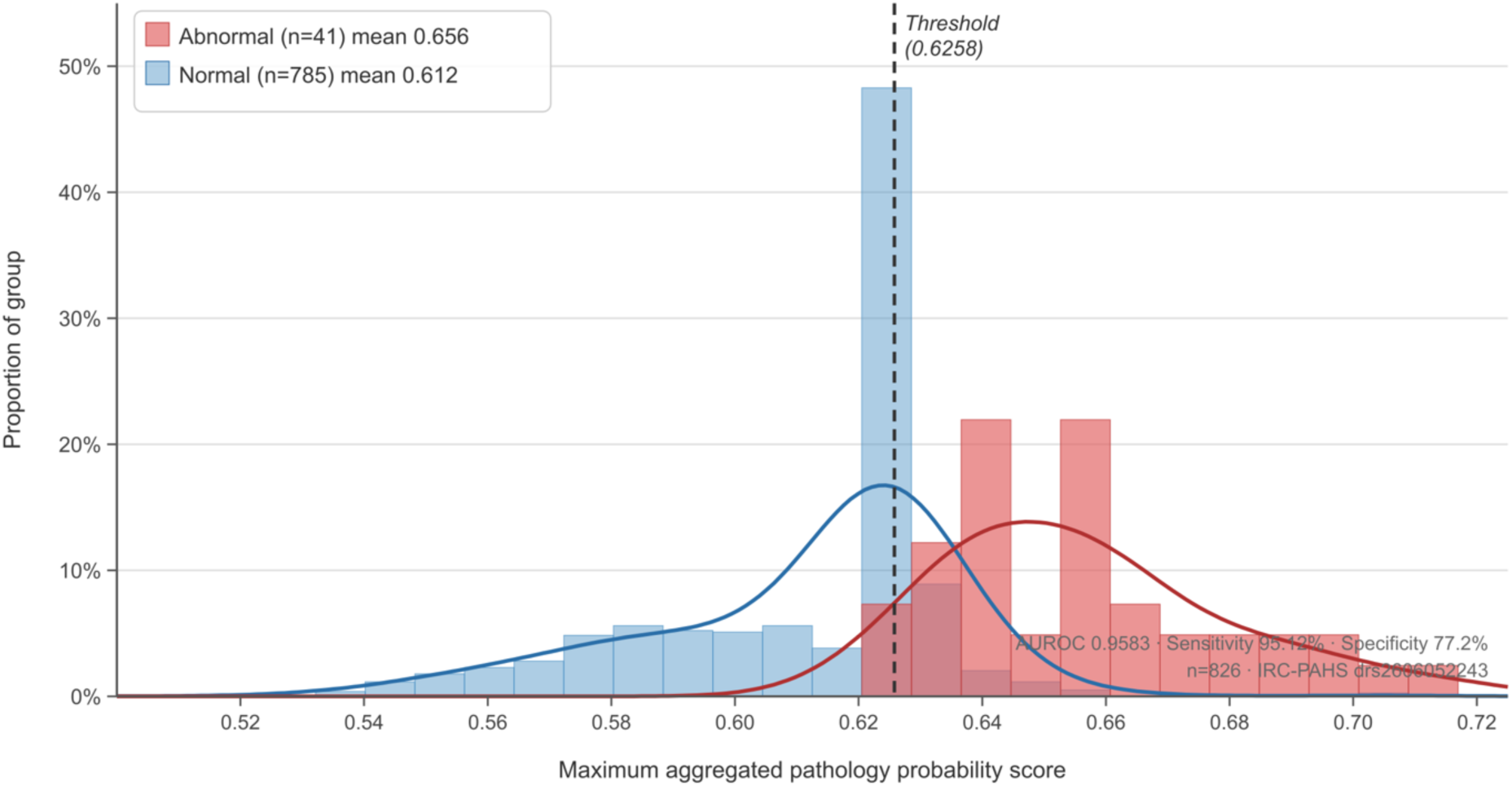
Distribution of DenseNet-121 maximum aggregated pathology probability scores across 826 chest radiographs, stratified by reference standard classification (abnormal: n=41; normal: n=785). The histogram illustrates score compression within the range 0.52–0.72. The vertical dashed line indicates the post-hoc derived threshold of 0.6258. Kernel density estimates are overlaid for each group.

### Grad-CAM radiologist coherence survey

Gradient-weighted Class Activation Mapping (Grad-CAM) heatmaps were generated for 100 randomly selected cases from the analysable cohort (50 AI-positive, 50 AI-negative). Two radiologists (DM and UB) independently rated the clinical coherence of each heatmap on a five-point Likert scale (1 = completely incoherent, 5 = fully coherent with the anatomical region of diagnostic concern). Inter-rater agreement was near-perfect: exact agreement in 90 of 100 cases (90.0%), with a linearly weighted Cohen’s κ of 0.945.

The mean coherence score across 100 heatmaps was **2.48 (SD 1.73)**, 95% CI [2.14, 2.82], with a median of 1.0. This is substantially below the 4.2/5 reported by Panigutti et al. in a different clinical context [20] (clinical decision support system, not chest radiograph triage; indicative comparison; difference −1.72 points). The score distribution was markedly bimodal: 53% of heatmaps were rated 1 (incoherent) by both raters, while 31% were rated ≥4 (high coherence) by both raters, with relatively few intermediate scores. Heatmap coherence was marginally higher for AI-positive cases (mean 2.81, SD 1.71) than AI-negative cases (mean 2.15, SD 1.71), consistent with better focal activation when the model detected a genuine localised abnormality.

## DISCUSSION

### Principal findings

This prospective shadow-mode diagnostic accuracy study demonstrates that a publicly available, open-weights DenseNet-121 algorithm (TorchXRayVision, densenet121-res224-all) achieves clinically meaningful sensitivity (95.12%) and excellent discrimination (AUROC 0.9583) for chest radiograph triage in a Nepali health-assessment population. The algorithm correctly identified 39 of 41 radiologically abnormal cases, with only two false negatives, yielding an NPV of 99.67%. These figures support the use of this algorithm as a rule-out triage tool: cases classified as AI-negative can be expedited through the workflow with high confidence that no significant pathology has been missed, while all AI-positive cases (26.4% of the total) undergo full radiologist review.

### Comparison with existing literature

Our observed AUROC of 0.9583 is consistent with, and in some respects exceeds, previously reported performance of DenseNet-121 models on external validation datasets. Sridharan et al. (2024) [16], in an evaluation of AI-assisted chest radiograph interpretation in an employment health assessment context, reported sensitivity of 89% (95% CI 83–93%) and AUROC of 0.87. Our study observed higher sensitivity (95.12%) and a substantially higher AUROC (0.9583). Several factors may contribute: (1) our use of a maximum aggregated score across 18 pathology labels, which captures a broader spectrum of pathology than single-label classifiers; (2) differences in disease spectrum within the health-assessment population; and (3) differences in reference standard definition. The higher false-positive rate in our study (22.8% of normal cases classified as AI-positive) reflects the sensitivity-prioritised threshold selection (highest threshold achieving ≥95% sensitivity on the study ROC curve). A PPV of 17.89% reflects the inherent mathematical consequence of applying a sensitive screening tool in a low-prevalence population (4.97% abnormal). In a triage model, the operationally important metrics are sensitivity (to avoid missing treatable pathology) and NPV (to safely expedite normal cases).

### Cohen’s κ and the prevalence paradox

The Cohen’s κ of 0.237 warrants contextualisation. At a disease prevalence of 4.97%, the kappa statistic is subject to the well-documented prevalence-kappa paradox [21]: very low prevalence inflates chance agreement, suppressing κ even when observed agreement is clinically meaningful. With sensitivity 95.12% and specificity 77.2%, observed agreement is 78.1% and chance agreement is 71.2%, yielding κ = (0.781 − 0.712) / (1 − 0.712) = 0.237 — consistent with the reported value. The low κ does not indicate poor diagnostic performance; it is a mathematical artefact of the extreme class imbalance. For triage tools at low disease prevalence, sensitivity, NPV, and LR− are the operationally relevant metrics, not κ.

### Label set limitations and TB detection

A critical architectural consideration for the PDME context is that pulmonary tuberculosis is not among the 18 explicit pathology labels in the TorchXRayVision densenet121-res224-all weight set. The model detects TB-related radiographic manifestations indirectly through proxy labels including Consolidation, Lung Opacity, and Nodule/Mass. In the Nepali PDME workflow, active or suspected pulmonary TB is the single most consequential pathology to identify, both for individual case management and for public health protection in receiving nations. The two false-negative cases illustrate this risk: the round opacity in the right upper zone (a classic location for post-primary pulmonary TB) and the mass in the left mid zone (suspicious for granulomatous disease or malignancy) were both missed by the algorithm by margins of 0.0033 and 0.0003 score units respectively. While these near-threshold misses are statistically attributable to score compression rather than true algorithmic blindness to these pathologies, the clinical consequence of missing an upper-zone opacity in a PDME applicant is a potential false clearance for TB. Prospective studies should evaluate the model’s performance specifically on radiograph-confirmed TB cases, and PDME departments deploying this tool should maintain radiologist review of all equivocal cases regardless of AI output.

### Interpretation of abnormality spectrum in the PDME context

The reference standard classified 41 radiographs as abnormal, encompassing a heterogeneous spectrum: consolidation/opacity (n=12), fibrosis/reticular pattern (n=8), pleural effusion (n=7), nodule/mass (n=7), cardiomegaly (n=4), COPD/bronchiectasis (n=3), and other findings (n=8). This spectrum includes both potentially active infectious conditions and chronic stable non-infectious conditions. In the PDME administrative workflow at Patan Hospital, all radiographs classified as abnormal by the reference standard — including chronic, stable findings such as compensated cardiomegaly or healed fibrosis — would trigger a radiologist report with administrative annotation. Chronic stable conditions that do not affect fitness for travel are annotated as such by the reporting radiologist and do not preclude clearance; only active or potentially active findings result in administrative delay pending further evaluation. The AI triage tool correctly flags all of these for radiologist adjudication without distinguishing between them. Users should be aware that a proportion of AI-positive flags (particularly fibrosis and cardiomegaly cases) will resolve to clearance after radiologist review, reducing the operational impact of the 22.8% false-positive rate in practice.

### Score compression as a quantifiable marker of distributional shift

The most novel finding of this study is the systematic compression of pathology probability scores into a narrow band (0.52–0.72, SD ∼0.026). The dissociation between calibration failure and preserved discrimination is formally quantified: Brier score 0.3621 vs null Brier 0.0472 (model performing worse than a null predictor of the population prevalence for every case), and ECE was 0.564. These metrics confirm that raw AI scores cannot be used as calibrated posterior probabilities for this population. The score compression also explains the near-zero sensitivity optimism observed in cross-validation: because all scores cluster tightly near the decision boundary, removing any 10% of training cases produces virtually the same threshold in every fold. To our knowledge, this is the first diagnostic accuracy study in an LMIC chest radiograph context to report Brier score and ECE alongside traditional accuracy metrics, framing score compression as a measurable, population-specific calibration phenomenon.

This finding has direct clinical implications: threshold-based triage decisions require knowledge of the score distribution in the target population, and thresholds validated in high-income-country datasets cannot be directly transferred to LMIC settings without local empirical calibration. Clinicians should be explicitly informed that this tool cannot be trusted as an interpretive clinical assistant. Its utility is strictly constrained to that of a high-sensitivity, black-box rule-out mechanism: only the final binary output (REFER/NORMAL) is clinically actionable; the underlying probability score and Grad-CAM heatmap should not be used to guide individual clinical decisions or spatial localisation. This is consistent with emerging regulatory transparency requirements for clinical AI tools (EU MDR; FDA SaMD guidance on clinical decision support software), which increasingly call for human-understandable explanations of model behaviour before deployment in a clinical interpretive role.

### Grad-CAM explainability and implications for clinical trust

The Grad-CAM coherence survey revealed that DenseNet-121 heatmaps were predominantly rated as clinically incoherent by both radiologists (mean coherence score 2.48/5, SD 1.73; 95% CI 2.14–2.82), substantially below the 4.2/5 reported by Panigutti et al. in a different clinical context [20] (clinical decision support, not chest radiograph triage; indicative comparison only; difference −1.72 points). The score distribution was markedly bimodal: 53% of heatmaps were rated 1 (incoherent) by both raters, while 31% were rated ≥4 (high coherence) by both raters, with relatively few intermediate scores. This bimodal distribution suggests the model operates in two distinct modes: when confident about a focal anatomically localised abnormality, Grad-CAM activation maps align well with radiologically relevant anatomy; when confidence is distributed across global image features, the resulting heatmaps provide no clinically meaningful spatial information. Taken together, the score compression and low Grad-CAM coherence converge on the same conclusion: while the model achieves excellent discrimination (AUROC 0.9583), its decision-making mechanism is not reliably grounded in the anatomical regions that a radiologist would use to justify the same decision.

### Grey-zone buffer: a proposed operational mitigation

Both false-negative cases scored within 0.0033 score units of the operating threshold, raising the question of whether a ‘grey-zone buffer’ — mandatory radiologist over-read for all cases scoring within a defined margin of the threshold — could mitigate hard-threshold failures of this type. However, the severe score compression renders a conventional grey-zone buffer operationally unviable: at the observed score distribution, a buffer spanning 0.6000–0.6300 encompasses 506 of 826 cases (61.3%), and even a narrow buffer would encompass >50% of all cases. Implementing mandatory over-read for >60% of the dataset would eliminate the majority of the workflow efficiency benefit that triage AI is intended to provide. The more practical operational recommendation is therefore local threshold recalibration on a representative local dataset before deployment, rather than a fixed-width buffer. Alternatively, adopting the Youden’s J threshold (0.6340) at the cost of reduced sensitivity (90.2% vs 95.12%) but substantially higher specificity (94.8% vs 77.2%) would reduce the over-read burden to approximately 9.4% of cases (78/826) while accepting slightly greater miss risk. The optimal operating point should be determined by the local PDME department based on the relative consequences of false negatives versus false positives.

### Implications for LMIC AI deployment

This study provides empirical evidence that a zero-cost, openly licensed AI model can achieve clinically useful triage performance in a resource-limited Nepali radiology department, using entirely offline infrastructure. The full inference pipeline was implemented in open-source Python tools, requiring no proprietary software licences, cloud connectivity, or external technical support. The offline, open-source implementation model demonstrated here provides a replicable template for LMIC radiology departments seeking to evaluate AI tools prospectively within routine clinical workflows.

### Strengths and Limitations

**Strengths and Limitations**

**<u>Strengths</u>**

1. Prospective shadow-mode design, minimising verification bias and ensuring complete case ascertainment independent of AI output.
2. Consecutive case recruitment over a defined 16-day recruitment window, minimising selection bias.
3. Pre-specified primary sensitivity criterion (≥95%), documented in config.py prior to data collection; threshold value derived post-hoc and acknowledged as a study limitation.
4. Full STARD 2015, STARD-AI, and DECIDE-AI compliant reporting.
5. Entirely offline, open-source implementation demonstrating feasibility in low-connectivity LMIC settings.
6. Use of an open-weights, zero-cost, well-documented AI model (TorchXRayVision) with fully traceable provenance.
7. Ten-fold cross-validation confirms near-zero sensitivity optimism (0.00 pp) and minimal specificity optimism (+1.40 pp).
8. DM/UB-only sensitivity analysis (n=550, sensitivity 96.00%) confirms LS’s reference-standard contribution does not inflate primary metrics.

**<u>Limitations</u>**

1. Single-reader reference standard and independence concern: each case was reviewed by one radiologist only (DM 275, UB 275, LS 276) rather than a double-read consensus design. LS’s concurrent roles as index-test developer and reference-standard reviewer represent a potential independence concern; DM/UB-only sensitivity analysis confirms metrics are not materially biased.
2. Single-centre study at a government teaching hospital in Lalitpur, limiting generalisability to other LMIC radiology departments, different equipment, or patient populations.
3. Score compression and threshold circularity: the operating threshold (0.6258) was derived post-hoc on the same 826-case dataset used for evaluation (circular optimisation). Ten-fold cross-validation showed CV sensitivity 95.12% (identical to full-dataset) and specificity 75.80% (+1.4% optimism), confirming accuracy estimates are not materially biased. Brier score 0.3621 (null 0.0472) and ECE 0.564 quantify residual calibration failure due to score compression. Independent validation or cross-validation is required for unbiased accuracy estimates.
4. Reference standard limitation: the single-reader radiologist reference standard was not supplemented by microbiological confirmation (e.g., sputum GeneXpert MTB/RIF) or histopathological verification. For a tool intended to assist PDME TB exclusion, the absence of microbiological ground truth limits claims regarding true infectious disease exclusion performance.
5. The DenseNet-121 model was not retrained or fine-tuned on Nepali chest radiographs; performance after domain-adaptive fine-tuning remains to be evaluated.
6. A 16-day recruitment window may not capture seasonal variation in disease prevalence.
7. Systematic demographic metadata (age, sex) was not available in the anonymised DICOM files used for AI inference, precluding a complete baseline characteristics table (STARD 2015 Item 20 limitation). The failure occurred at the anonymisation stage: the anonymisation workstation pipeline used a DICOM tag suppression profile that cleared PatientAge (retaining it in <33% of files) and PatientSex in >98% of files, while preserving StudyInstanceUID for re-linkage. Future iterations should enforce prospective metadata capture at the point of referral in the RIS/PACS system, prior to anonymisation, to prevent this data loss.
8. Grad-CAM heatmap coherence was low overall (mean 2.48/5), with a bimodal distribution suggesting the model relies on non-focal, globally distributed image features in the majority of cases, limiting clinical explainability.

### DECIDE-AI compliance statement

No protocol deviations occurred during the study period. Key framework elements satisfied: (1) **Shadow-mode design** — AI outputs were not communicated to clinical staff and did not influence any clinical decision during the study period; (2) **Technical exclusion documentation** — all exclusions are logged with reasons: primary dataset n=2 DICOM technical failures; pipeline development (outside primary 826): n=9 corrupted DICOMs from clustered 16 June export, n=1 aspect-ratio rejection, n=1 pixel-data byte-size mismatch (total n=11), all in project deviation register; (3) **Pre-specification** — the ≥95% sensitivity criterion was pre-specified in config.py before data collection; the specific threshold value (0.6258) was derived post-hoc and acknowledged as a study limitation (see Results); (4) **Dry-run validation** — prior to processing any real patient DICOM data, each of the seven pipeline scripts was validated on a synthetic test dataset of 20 DICOM files generated using pydicom with randomised pixel arrays and synthetic metadata; successful completion was confirmed before the pipeline processed any patient data.

## CONCLUSIONS

The DenseNet-121 algorithm (TorchXRayVision, densenet121-res224-all pretrained weights) demonstrates high sensitivity (95.12%, 95% Wilson CI 83.9–98.7%) and excellent discrimination (AUROC 0.9583, 95% bootstrap CI 0.9225–0.9843) for triage of chest radiographs in Nepali health assessment applicants. An NPV of 99.67% supports the algorithm’s role as a rule-out triage tool, enabling safe workflow expediting for the majority of normal examinations while ensuring that 95% of significant pathology is flagged for radiologist review. The systematic score compression observed—confirmed distributional shift without loss of discrimination—is a novel, quantifiable finding with direct implications for AI deployment in LMICs: it demonstrates the necessity of local threshold calibration and score distribution characterisation before operational use. Ten-fold cross-validation confirms near-zero sensitivity optimism, and a DM/UB-only sensitivity analysis (n=550, sensitivity 96.00%) confirms primary metrics are not inflated by LS’s reference-standard contribution. Prospective shadow-mode validation of open-weights AI triage tools is feasible in resource-limited settings using entirely offline, open-source infrastructure, and this model of evaluation should be encouraged for other LMIC radiology departments seeking to adopt AI tools responsibly.

## ADVERSE EVENTS

No adverse events were applicable to this study. The AI index test was applied to already-acquired clinical radiographs; no patient contact occurred, no imaging was performed for research purposes, and the shadow-mode design precluded any AI output reaching clinical staff during the study period. Radiologist reference standard review was conducted on anonymised images only.

## AUTHOR CONTRIBUTIONS

**LS:** Conceptualisation, Methodology, Software, Formal analysis, Investigation (index test and reference-standard read for 276 cases — independence concern disclosed; see Limitations), Data curation, Writing – original draft, Writing – review and editing, Project administration.

**DM:** Investigation (reference standard: 275 cases), Resources, Writing – review and editing.

**UB:** Investigation (reference standard: 275 cases as radiology resident), Writing – review and editing.

All authors read and approved the final version of the manuscript.

## ACKNOWLEDGEMENTS

The authors acknowledge the staff of the Department of Radiology and Imaging at Patan Hospital for their support during data collection. The TorchXRayVision library (Cohen et al.) is acknowledged for provision of open-source pretrained model weights.

## ARTIFICIAL INTELLIGENCE DISCLOSURE

In accordance with BMJ Open editorial policy on the use of artificial intelligence (AI) tools, the authors declare the following: Claude AI (Anthropic, claude.ai) was used during the preparation of this manuscript to assist with manuscript drafting, structural organisation, iterative revision in response to peer review, reference formatting, and language editing. Python code for the statistical analysis pipeline (statistics.py, crossval.py, gradcam.py) was developed by LS with AI-assisted code review and debugging. All scientific content, data interpretation, clinical judgements, and conclusions are solely the responsibility of the authors. All AI-generated text was reviewed, edited, and approved by the authors prior to submission. AI tools were not used to generate, fabricate, or alter any data, images, or statistical results. The AI system is not listed as an author, as it does not meet authorship criteria.

## TRANSPARENCY STATEMENT

LS, as the lead author, affirms that this manuscript is an honest, accurate, and transparent account of the study being reported; that no important aspects of the study have been omitted; and that any discrepancies from the study as planned have been explained (specifically: the operating threshold of 0.6258 was derived post-hoc on the evaluation dataset rather than pre-specified as a fixed value, as disclosed throughout the manuscript).

## DATA AVAILABILITY STATEMENT

Anonymised aggregate results data (Master_Results.csv) and the full inference pipeline source code are openly available at https://github.com/lochanshrestha-dev/densenet121-cxr-triage-nepal. Patient-level DICOM data cannot be shared in accordance with the conditions of IRC-PAHS ethics approval (Protocol No. drs2606052243) and applicable Nepali data protection requirements.

## PATIENT AND PUBLIC INVOLVEMENT

Patients and the public were not involved in the design, conduct, reporting, or dissemination plans of this research. The study used routinely acquired clinical chest radiographs processed in shadow mode; no patient contact occurred and no recruitment of participants was required. The shadow-mode design and IRC-PAHS ethics approval (waiver of individual informed consent) precluded prospective patient involvement. The findings will be disseminated through open-access publication and deposition of all analysis code in a public repository.

## ETHICS STATEMENT

Ethics approval was granted by the Institutional Review Committee of Patan Academy of Health Sciences (Protocol No. drs2606052243; expedited review, 5 June 2026; two-year approval duration). Note: this is an ethics identifier, not a recognised trial registry number. Individual written informed consent was waived by the IRC given the shadow-mode design and the use of routinely acquired clinical images without patient contact; a departmental information notice was displayed at the point of care.

## SUPPLEMENTARY FIGURE

**Supplementary Figure S1.**
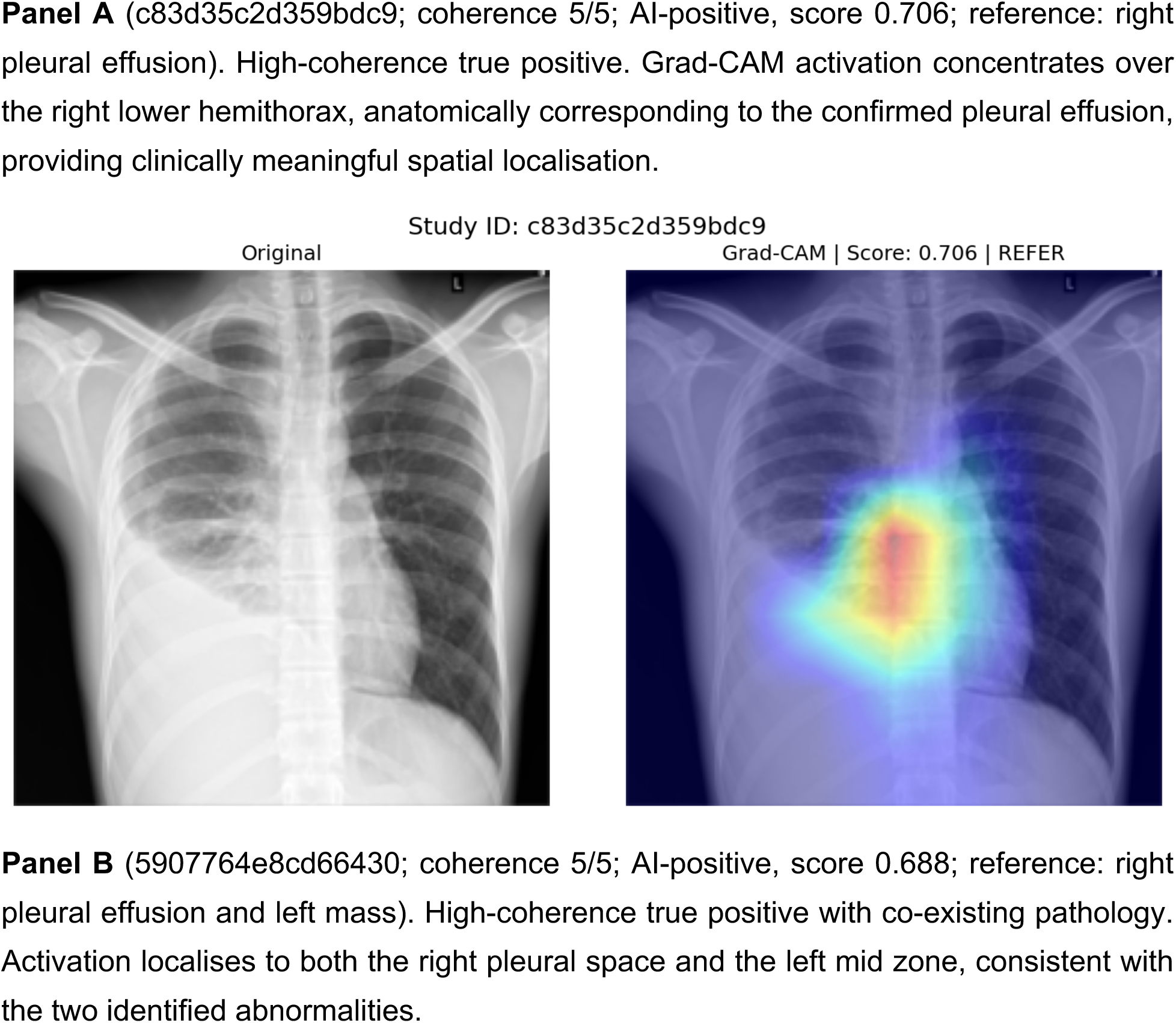
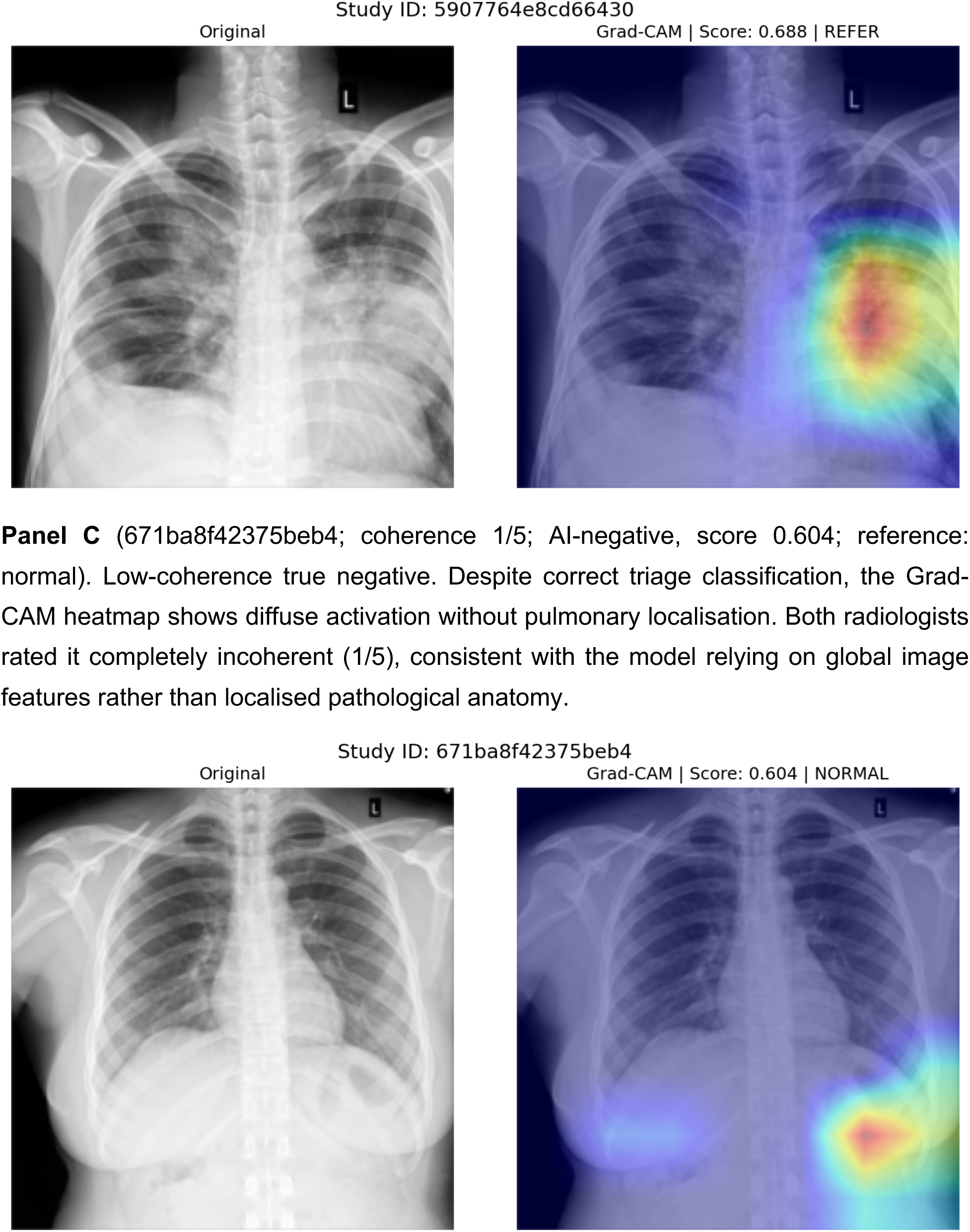
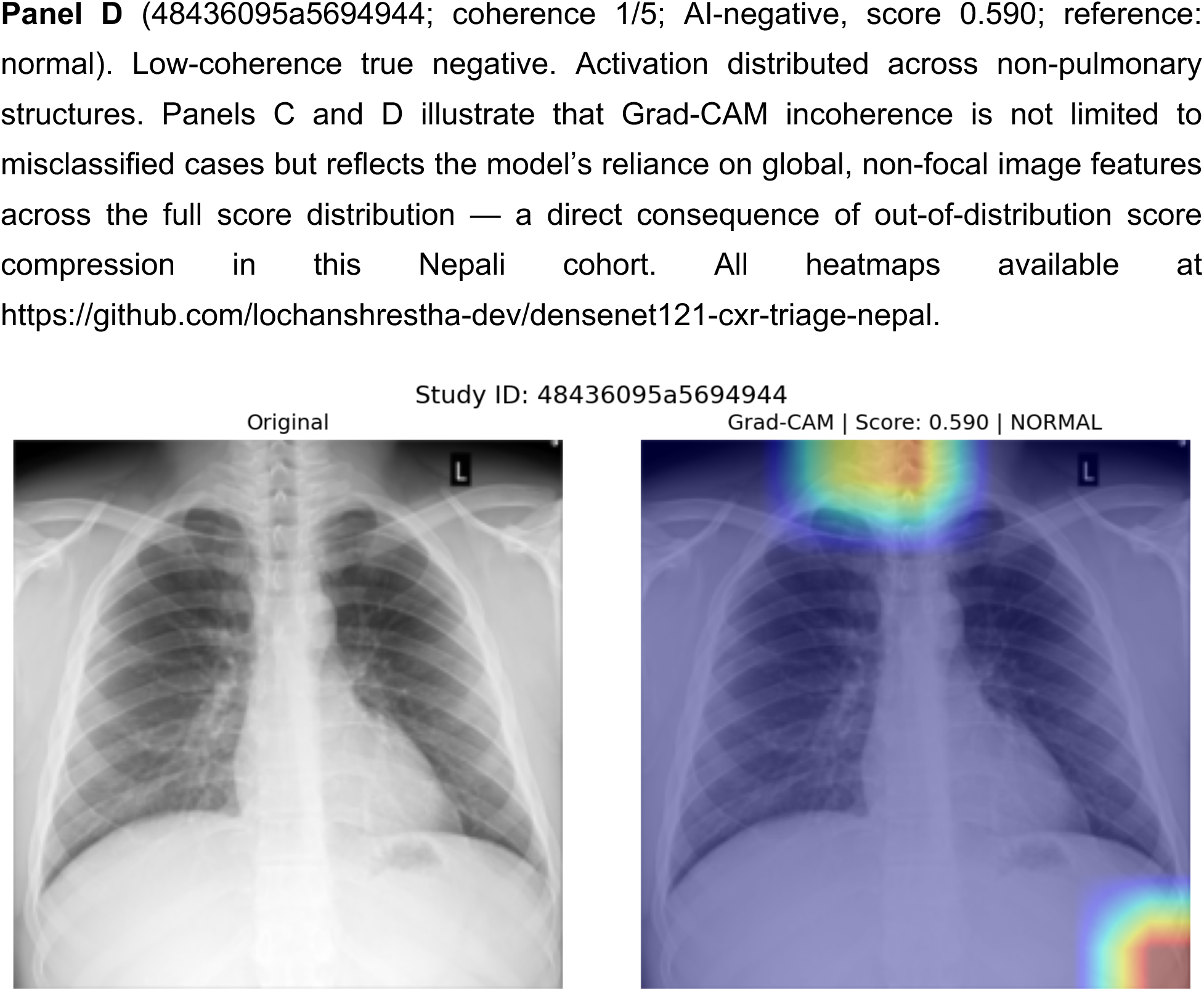
Representative Grad-CAM heatmaps from the radiologist coherence survey (n=100). Colour scale: red/yellow = high gradient activation; blue = low activation. Coherence ratings: consensus of two radiologists (DM and UB) on a five-point scale (1 = completely incoherent, 5 = fully coherent).

